# Cost savings from managing hypertension in primary health care clinics in Kuwait

**DOI:** 10.1101/2021.05.07.21256767

**Authors:** Ayah Odeh, Syed Mohamed Aljunid, Rihab Al-Wotayan, Mahmoud Annaka, Mohammad Al Mari

## Abstract

**Background:** Generic medications are one of the most common solutions for bringing down pharmaceutical costs for both patients and health care providers. Efforts to increase uptake of generics include policies to support generic substitution and prescription. The aim of this study is to estimate the total cost of drugs prescribed by physicians in selected primary health care centers for managing hypertension and the potential cost savings from substituting generic drugs for branded ones

**Methods and Findings:** One thousand patients with hypertension were randomly selected from the Primary Care Information System database from among patients who sought treatment at three primary health care centers from Al-Jahra governorate in Kuwait from January to December 2018. Generic antihypertensive drugs were substituted for branded ones, and cost savings were calculated by referring to the International Drug Price Indicator Guide. The mean age of 1,000 patients was 57.01 (SD = 11.82) years. Most (57.4%) of the patients were females, and 50.2% were Kuwaitis. The mean number of drugs per prescription was 1.78 (SD = 1.25; range: 1 to 9 drugs). The total number of drugs prescribed was 1,781, with a total cost of KD 10,093 and with a mean of KD 10.09 (SD = 7.34). Only 71 generic drugs had been prescribed, making the generic prescription rate 4.0%. The total number of antihypertensive drugs prescribed was 1,206 (mean: 1.21; SD = 0.46), with a cost of KD 7,678.5 (mean = KD 7.68; SD = 4.06) and with ACE inhibitors as the most prescribed class at 32.19%. Eight hundred ninety antihypertensive drugs were substituted for 774 patients at estimated cost savings of KD 5,675; that is, substituting generic drugs reduced antihypertensive drug cost by 74%.

**Conclusion:** Generic drug prescription appears to be low among primary care physicians in health care centers in Kuwait, but these centers could see substantial cost savings from substituting generic antihypertensive drugs for branded ones. Active interventions are needed to encourage generic prescription among health practitioners to reduce the overall pharmaceutical expenditures.

## Introduction

Countries around the world continue to grapple with increasing health care expenditures, an essential component of which are drugs [1, 2, 3] with the rise in health care costs driven mainly by population growth, increasing life expectancies, and new technologies. Spending on health accounts for more than 10% of gross domestic product of most developed countries, with drugs contributing to no less than a fifth of health expenditures [4]. Pharmaceutical spending is growing at an average of 5.7% every year, faster than other components of health expenditures [5], and the overwhelming spread of chronic diseases is one of the leading causes of the rising health care costs. Patients with chronic illnesses utilize more services and increase the demand for pharmaceuticals [6, 7, 8]. For these reasons, it is imperative that policy makers act innovatively to control these ever-growing expenses without jeopardizing the quality of care provisioned to patients.

Generic drugs are the most common solution for curbing pharmaceutical expenditures and bringing down costs for both patients and governments. They offer the same therapeutic benefit but at prices up to 80% less than the costs of originators [9]. Encouraging their uptake through generic prescription and substitution has become an integral part of health care policies [10, 11, 12]. The United States achieved a generic drug prescription rate (GDPR) of 90% that generated savings of up to $265.1 billion [13], and the United Kingdom similarly succeeded in achieving a generic prescription rate of 84%, with overall savings estimated to be £7.1 billion since 1976 attributable to generic prescription [14]. Furthermore, in Canada, even though generic drugs represent 73% of all drugs, they account for less than 20% of the total costs spent on drugs due to their low price [15]. In Kuwait, studies on generic prescription rates are limited. One of the studies conducted in 2010 reported that the generic prescription rate was 17.7% [16], a percentage deemed extremely low by WHO standards [17].

Meanwhile, as a lifelong illness, hypertension exerts a significant economic burden, especially in developing countries. In the coming years, high blood pressure might cost nearly $1 trillion globally in direct health spending annually and $3.6 trillion in indirect costs [18]. As with other chronic conditions, patients with hypertension rely on health services and drugs for lifelong treatment. In Kuwait, hypertension is one of the most debilitating diseases, ranking as the sixth leading cause of death in the country at 37% of all deaths [19]. With 27.8% of Kuwait’s population estimated to be suffering from hypertension that requires prolonged treatment [19], reducing the cost of antihypertensive drugs could translate to remarkable savings during patients’ lifetime. Hence, in this study, we sought to determine the rate of generic drug prescription among primary health care physicians in Kuwait and to estimate the total cost of drugs prescribed for hypertension management as well as the potential cost savings from substituting generic antihypertensive drugs for branded ones.

## Methods

### Study design, sampling determination, and source of data

We conducted this cross-sectional study in three primary health care clinics (PHCCs) located in Al-Jahra governorate in Kuwait. We searched the Primary Care Information System database of patients using the inclusion criteria of a diagnosis with hypertension and prescriptions for antihypertensive drugs from January to December 2018. Excluding patients without hypertension or with incomplete drug information, we selected the first 1,000 patients as the sample for this study after we randomly assigned them using Microsoft Excel’s random number function and arranged the numbers in descending order. We used the previously reported PHCC GDPR for Kuwait of 17.7% [16] to estimate the sample size and used OPENEPI to calculate that a minimum of 224 patients were required, but we selected a sample of 1,000 patients to compensate for the lower GDPR in the benchmarked study. We also gathered the patients’ demographic data, diagnoses, and drugs from the Primary Care Information System, ascertaining what drugs had been chosen for substitution from the list of antihypertensive agents in the *2017 Report of the American College of Cardiology/American Heart Association Task Force on Clinical Practice Guideline for the Prevention, Detection, Evaluation, and Management of High Blood Pressure in Adults*.

### Data management procedures

We obtained public prescription prices from the Drug Price List for January 2020 released by the Kuwaiti Ministry of Health’s (MOH) Pharmaceutical & Herbal Medicines Registration & Control Administration, excluding drugs and other pharmaceutical preparations listed without prices. To calculate the cost of antihypertensive drugs for our analyses, we standardized the treatment duration to one month. We calculated the defined daily dose based on the usual adult maintenance dose for managing hypertension [20]. Hence, the price for one month was as follows: defined daily dose (DDD) × 30 days × unit price = price in KD/month/drug [21]. For non-antihypertensive drugs, we assumed the cost of one pack except for Vitamin D3 50,000 IU, the cost of which we calculated based on the dosage of one unit per week for a month. We reviewed all prescriptions to identify drugs that had been prescribed as generic or by brand name to calculate the GDPR, which we calculated by dividing the number of generic drugs prescribed by the total number of drugs prescribed.

For the substitutions of generic antihypertensive drugs, we used prices from the *International Drug Price Indicator Guide 2015*, released by the Management Sciences for Health, Inc., in collaboration with the WHO [22] using the median buyer price/unit. We used the same method to calculate the costs of the originally prescribed drugs along with factoring in shipping costs, inflation rate, and USD conversion rate: DDD × median unit price from guide × 30 days × 10% shipping × inflation rate × conversion rate = price in KD/month/generic drug. The guide provides generic prices only for essential medications, and as such, we could not substitute all antihypertensive drugs that the physicians at the three PHCCs that we studied had prescribed for their patients. Ultimately, we imputed three main costs for analysis: the total cost of the prescriptions, the cost of antihypertensive drugs, and the amount of savings generated by generic substitution.

### Statistical analysis

We analyzed the study data using Statistical Package for Social Sciences, version 24, with significance set at 5%. Categorical variables were summarized using frequencies and proportions, and continuous variables were summarized using means and standard deviation, and medians and interquartile ranges. We also used the Mann–Whitney U test to compare the medians of the total prescription costs before and after generic substitution.

### Ethical Approval

The study was approved by the Research and Ethics Committee in the Health Sciences Center, Kuwait University and the Standing Committee for coordination of health and medical research in the Ministry of Health.

## Results

### Patient demographics

In a random sample of 1,000 patients with hypertension, more than half of patients (57.4%) were female, and there were slightly more Kuwaiti (50.2%) than non-Kuwaiti (49.8%) patients. The mean age of patients was 57.01 years (SD = 11.82), and the median was 56 years. Most of the patients were in the age group of 45 to 64 (61.2%), and elderly patients (65 and above) represented only 24.5% of the sample. The duration of illness was only available for 461 patients and had a mean of 11.11 (SD = 5.8) years (Table 1).

**Table 1.** Clinical and Sociodemographic Profile of Patients.

| Characteristic | Number of patients (%) |
| --- | --- |
| Gender |  |
| Female | 574 (57.4%) |
| Male | 426 (42.6%) |
| Age (mean ± SD) | 57.01 ± 11.82 |
| Below 45 years | 143 (14.3%) |
| 45–64 years | 612 (61.2%) |
| 65 and above | 245 (24.5%) |
| Nationality |  |
| Kuwaiti | 502 (50.2%) |
| Non-Kuwaiti | 498 (49.8%) |
| Duration of illness |  |
| Less than 5 years | 79 (17.1%) |
| 5–9 years | 106 (23.0%) |
| More than 9 years | 276 (59.9%) |
| Total | 461 (100%) |
| Mean 11.11 ± 5.81 | |

### Drug prescriptions

The total number of drugs prescribed for the 1,000 patients was 1,781, with a mean number of drugs prescribed per prescription of 1.78 (SD = 1.25) and a median of 1. Over half (60.4%) of the patients had been prescribed one drug per visit. Table 2 presents the data on drugs distributed to patients in this study.

**Table 2.**
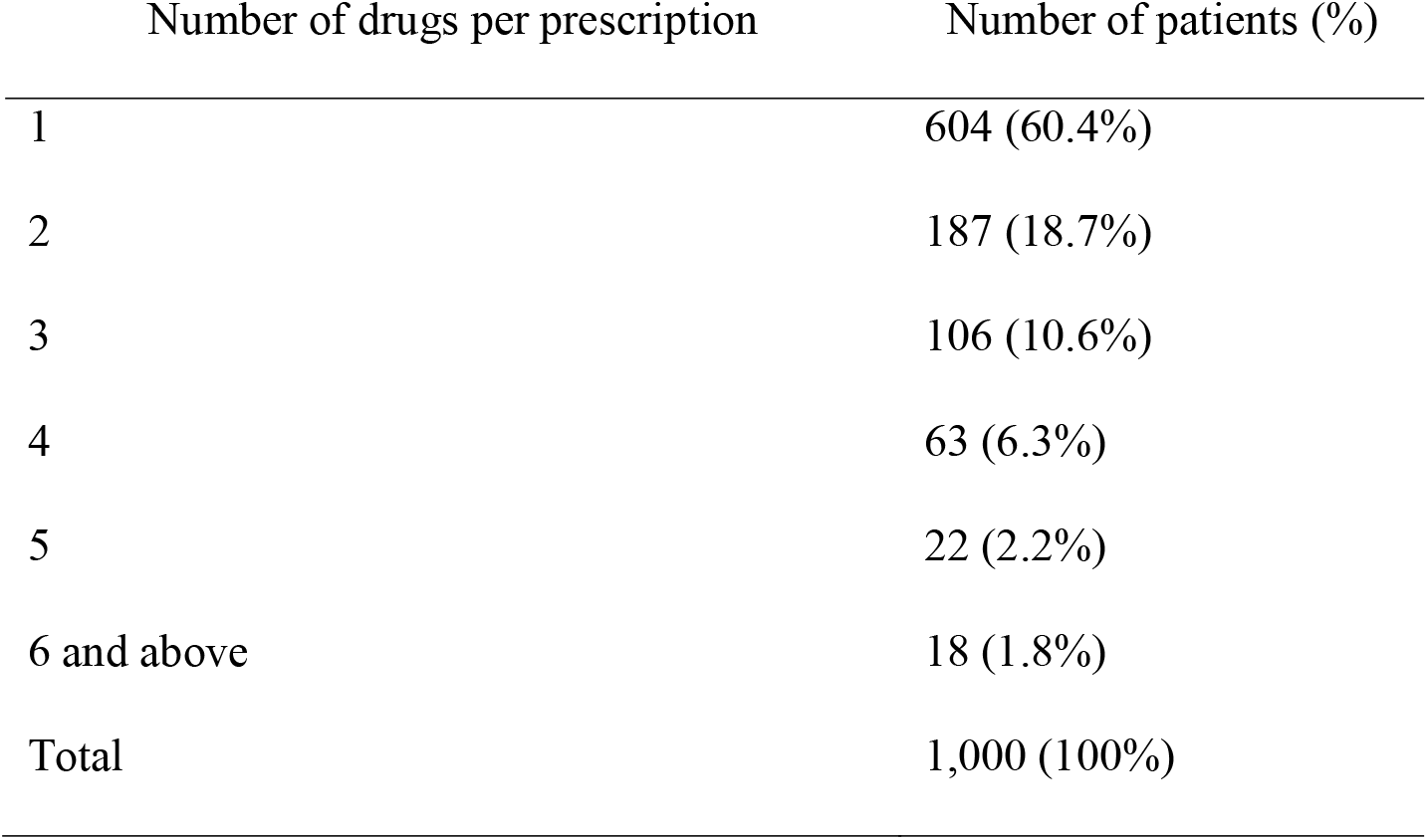
Distribution of Drugs Prescribed to Patients.

Providers at the three PHCCs in this study prescribed a total of 71 generic drugs to 56 patients, making the generic prescription rate 4.0%. The average number of generic drugs was 0.07 (SD = 0.32), with a median of 0, whereas the average number of branded drugs was 1.71 (SD = 1.14), with a median of 1 (Table 3).

**Table 3.** Distribution of Branded and Generic Drugs.

| Type of drug prescribed | Number of drugs (%) |
| --- | --- |
| Generic | 71 (4.0%) |
| Branded | 1,710 (96.0%) |
| Total | 1,781 (100%) |

The number of antihypertensive drugs prescribed in this study was 1,206, averaging 1.21 (±0.46) per patient, with a median of 1. Most patients (81.7%) had been prescribed only one antihypertensive drug (Table 4).

**Table 4.** Distribution of Antihypertensive Drugs among Patients.

| Number of antihypertensive drugs | Number of patients (%) |
| --- | --- |
| 1 | 817 (81.7%) |
| 2 | 160 (16.0%) |
| 3 | 23 (2.3%) |
| Total | 1,000 (100%) |

The most prescribed class of antihypertensive drugs was angiotensin-converting enzyme inhibitors (ACEIs), representing 32.2%, followed by calcium channel blockers (CCBs), beta blockers (BBs), and angiotensin ii receptor blockers (ARBs). Tables 5 and 6 present the generic antihypertensive drugs prescribed by type and the branded antihypertensive drugs prescribed, respectively.

**Table 5.** List of Generic Antihypertensive Drug Types Prescribed.

| Antihypertensive drugs prescribed by brand name | Number (%) |
| --- | --- |
| Alpha-blockers & receptor agonists | 10 (0.8%) |
| ACEIs | 385 (32.2%) |
| ARBs | 126 (10.6%) |
| BBs | 136 (11.4%) |
| CCBs | 182 (15.2%) |
| Centrally acting antiadrenergic | 59 (4.9%) |
| Combination drugs | 254 (22.0%) |
| Diuretics | 44 (4.0%) |
| Total | 1,196 (100%) |

**Table 6.** List of Branded Antihypertensive Drugs Prescribed.

| Antihypertensive drugs prescribed by generic<br>name | Number (%) |
| --- | --- |
| Amlodipine | 1 (10%) |
| Captopril | 2 (20%) |
| Cilazapril | 3 (30%) |
| Clindipine | 1 (10%) |
| Losartan | 1 (10%) |
| Valsartan | 2 (20%) |
| Total | 10 (100%) |

### Drug costs

The overall cost of drugs for all 1,000 patients was KD 10,092.99, with a mean of KD 10.09 (SD = 7.34) and a median of KD 8.06. Most patients’ prescriptions (83%) cost less than KD 15. Tables 7 and 8 present the data on the overall and hypertensive drug costs and the distributions among the patients.

**Table 7.** Drug Costs for 1,000 Patients.

| Cost | Total (KD) | Mean | SD | Median | IQR | Mode |
| --- | --- | --- | --- | --- | --- | --- |
| Total cost of prescription | 10,092.99 | 10.09 | 7.34 | 8.06 | 6.38 | 8.06 |
| Cost of antihypertensive drugs | 7,678.49 | 7.68 | 4.06 | 7.16 | 5.73 | 11.76 |
| Cost of prescription after generic substitution | 4,417.97 | 4.42 | 6.53 | 2.13 | 3.24 | 0.56 |
| Cost of antihypertensive drugs after generic substitution | 2,003.47 | 2.00 | 2.26 | 0.91 | 3.24 | 0.56 |
| Amount of cost saving | 5,675.04 | 5.68 | 4.32 | 5.19 | 7.81 | 0 |

**Table 8.** Drug Cost Distribution among Patients.

| Cost (KD) | Number of patients (%) |  |  |
| --- | --- | --- | --- |
|  | Cost of prescription | Cost of antihypertensive drugs | Amount of savings |
| Less than 5.00 | 247 (24.7%) | 312 (31.2%) | 227 (22.7%) |
| 5.00–9.99 | 364 (36.4%) | 402 (40.2%) | 232 (23.2%) |
| 10.00–14.99 | 219 (21.9%) | 244 (24.4%) | 294 (29.4%) |
| 15.00 and above | 170 (17%) | 42 (4.2%) | 247 (24.7%) |
| Total | 1,000 (100%) | 1,000 (100%) | 1,000 (100%) |

### Cost of antihypertensive drugs

The total cost of antihypertensive drugs was KD 7,678.49, representing 76% of the total cost of all prescriptions. The average cost of antihypertensive drugs was KD 7.68, and more than a third (40.2%) of the antihypertensive drugs cost less than KD 10.

### Amount of cost savings

The total amount of savings achieved by substituting generic antihypertensive drugs for branded drugs for 774 patients was KD 5,675.04. Substituting 890 antihypertensive drugs (73.8%) reduced total prescription costs by 56.2% and total antihypertensive drug costs by 73.9%.

### National cost savings

The prevalence of hypertension in Kuwait is estimated to be 27.8% [19]. On the basis of this prevalence, we attempted to calculate national savings from substituting 22 types of antihypertensive drugs in Kuwait. Table 9 presents our findings.

**Table 9.** National Savings from Generic Substitution of Generic Antihypertensive Drugs.

|  |  |
| --- | --- |
| Prevalence of hypertension | 27.8% |
| Population of Kuwait in 2019 | 4.7 million |
| Median amount of saving/month/patient | 5.187 |
| Number of people with hypertension | $4.7 \text{ million} \times 27.8\% = 1,306,600$ |
| Amount saved for hypertension population/year | $1.3 \text{ million} \times 5.187 \times 12 = \text{KD } 81,328,010$ |

To check the robustness of the results, we conducted a sensitivity analysis. We calculated the savings for prevalence rates 10% lower and 10% higher than the actual prevalence and for generic substitution rates 26% above and below the actual substitution rates, representing the best and worst-case scenarios respectively. Table 10 presents the best- and worst-case scenario sensitivity analysis findings.

**Table 10.** National Savings from Substituting Generic Hypertensive Drugs for Brand Name.

| Component | Savings (KD) |
| --- | --- |
| 17.8% prevalence and 100% substituted (Best case) | 70,274,400 |
| **27.8% prevalence and 74% substituted (Base)** | **81,328,010** |
| 37.8% prevalence and 48% substituted (Worst case) | 71,739,108 |

## Discussion

The main findings of this study are a low GDPR and a promising amount of savings achieved by substituting generic antihypertensive drugs for branded drugs. The majority of the patients were below 64 years of age, even though previous researchers have well established that hypertension prevalence in Kuwait [23] and worldwide increases with age, especially after 60 [24]. However, these numbers could merely reflect the governorate’s age distribution: in recent statistics, elderly citizens above age 65 represented only 2.1% of Al-Jahra’s inhabitants and 98% were below 64 years [25]. In 1980, hypertension prevalence among Kuwaiti nationals and non-nationals was 8.3% in men and 12.9% in women, and in 1999, the prevalence among Kuwaitis only was reported to be 28.3% in men and 22.9% in women [23]. In this study, the sample had more female than male patients, and in a previous study, on drug compliance among patients with hypertension visiting primary health care clinics, female participants also outnumbered males (59.8% to 40.2%) [26].

The GDPR in this study was 4.0%, lower than the rate of 14.7% previously reported in Al-Jahra governorate primary health care clinics, and the median number of drugs per prescription was also lower than the previously reported median of 2.6 [16]. These differences, however, could be attributed to the different methodologies in selecting patients. The previous study’s authors included prescriptions for general illnesses but excluded chronic diseases such as hypertension, whereas in this study, we studied only patients with hypertension. The objectives of the two studies were different, hence the different inclusion criteria and accompanying differing results.

That being said, both the GDPRs reported in Kuwait in 2010 and in this study are low by WHO standards. Ideally, 100% of prescriptions are for generic versions [17]. The rates in the neighboring Gulf countries of Kingdom of Saudi Arabia, Bahrain, and United Arab Emirates (2.9%–19.5%, 14.3%, and 7.4%, respectively) are similarly low [16]. In the UK, the GDPR is 84% [14], and in the United States, it is nearly 90% [13].

In 2010, the low generic prescription rate was attributed to the feature of the primary health care electronic system that it only showed drugs by their brand names [16]. This system has since been updated to include both generic international non-propriety and brand names of drugs, and even with that change, prescription by generic names actually decreased. Until recently, the MOH had no official requirement that physicians prescribe generic drugs. Not until January 25, 2020, did a new ministerial decree oblige physicians to prescribe medications by their generic non-propriety names [27].

Furthermore, physicians in Kuwait do not receive adequate training on hospital supply management, despite their having a key role in selecting treatments. Instead, they base their decisions regarding pharmaceuticals on personal opinion, training, and relationships formed with pharmaceutical companies through drug company representatives and sponsored conventions but without regard to cost. In turn, drug companies encourage their representatives to build strong relationships with physicians through value-added services to influence their prescription [28]. Moreover, because physicians often encounter brand names of drugs during their training, they are more likely to remember these drugs [16]. Therefore, educating physicians on the basics of health care supply management, such as informing them of prices and the potential savings from substituting generics, can help in promoting generic drug prescription in Kuwait.

Moreover, the MOH should have physicians receive their medication information from academics rather than medical company representatives. Researchers established that in contrast to detailing by medical representatives, academic detailing had a positive influence on physicians’ prescription habits that had relied on presenting neutral and evidence-based drug information [29]. The ministry could organize regular interactive sessions led by experts to supply physicians with information on prescription drugs uses.

In addition, although the MOH has the unofficial policy of allowing pharmacists to switch drug brands according to their availability, the availability of generics in Kuwait is limited, restricting pharmacists’ ability to substitute [30]. In this study, nearly all of the antihypertensive drugs prescribed had been branded ones, and the generic drugs prescribed were mainly vitamins, supplements, and antibiotics, but drugs used to treat chronic diseases were prescribed mostly by brand name, perhaps reflecting drug companies’ marketing focus on the chronic “diseases of the century” and the drugs to treat them that hold the biggest share of the pharmaceutical market. Furthermore, the five most-prescribed antihypertensive drugs by brand name were all originator-branded drugs that have generic versions on international markets and were among the highest-costing drugs, indicating an opportunity for cost saving by substituting generics.

Kuwait has a high per capita income, which means the demand for quality in health care is high, but the limited indigenous manufacturing capabilities of pharmaceuticals led to dependence on imported pharmaceuticals. In 2017, originator drugs led the pharmaceutical market sales in Kuwait, and generic drug market penetration was limited [31]. In 2019, generics made up only 21.6% of the Kuwaiti pharmaceutical market, in contrast to UK and USA where generic prescriptions are as high as 89% and 81%, respectively [28]. People usually equate high cost with high quality, which has popularized more expensive branded medications over generics and reduced demand for them. Additionally, the high per capita income of most of the Kuwaiti population has led to low patient acceptance of generic drugs [32]. Patients with chronic diseases are accustomed to taking more expensive branded drugs, and they might refuse generic versions for a variety of reasons such as misconceptions regarding their quality. For instance, authors of one study found conflicting opinions on generic and branded medications among patients with diabetes in Kuwait, with some believing that generics were less effective and had more side effects than foreign brands [33]. In another study, in countries where reliance on the public health sector is predominant, patients are more likely to demand the costliest treatments. Patients are aware that generic drugs cost less, but they demand the branded versions because they do not bear the costs [28], which raises the issue of educating patients on the bioequivalence of generics to increase their acceptability. New terms and specifications for the purchase and sale of generic medications in Kuwait were approved back in October 2019, and the MOH has also decided that generic medications will constitute 20% of all medications in Kuwait and branded ones will represent 80% [34,35].

During the time period that we studied (January to December 2018), most patients with hypertension had been prescribed one antihypertensive drug, and only 2.3% had been prescribed three drugs. In the Middle East, 40.5% of patients received two drugs for hypertension, and only 23.5% were on monotherapy [36]. In one study in Saudi Arabia, 48.9% of patients received one antihypertensive drug, 31% received two, and 23.3% received three or more [37], and results were similar in a study in Bahrain. Most patients in the latter study (62.9%) were treated with monotherapy [38]. In Jordan, 46.2% of patients with hypertension in one study received one drug for managing hypertension, and only 15.6% received three or more drugs [39]. By contrast, in the United States, most patients with hypertension receive multiple drugs to achieve better control [40].

In this study, angiotensin-converting enzyme inhibitors were the most prescribed class of antihypertensive agents, and of the drugs prescribed, the originator-brand ACEIs and ARBs were among the costliest. In 1999, the Central Medical Stores had BBs as the top-consumed drugs at 44.7%, followed by ACEIs at 16.9%, centrally acting agents at 13.3%, CCBs at 12.5%, diuretics at 12%, vasodilators at 0.3%, and alpha-blockers at 0.2% [23]. These numbers reflected the whole country’s consumption, including primary health care clinics, although ARBs were just newly introduced in Kuwait and as such were not included in the calculations [23]. In previous years, ACEI prescription increased, and indeed, ACEIs were the most prescribed in this study (32.19%). Among the ACEIs, lisinopril was the most prescribed (37.2%), which was also the case in 1999 [23]. Lisinopril is also the most widely used ACEI in the United States [40].

In this study, ACEIs accounted for almost half of the total prescription cost, and the major ACEIs prescribed were all originator drugs. Gradual shifts to the use of generic drugs could begin with ACEIs given that they are prescribed the most and cost the most. The second category worth considering for substitution could be CCB. Analysis of the combination drugs revealed that ARBs were prescribed more than CCBs but that CCBs represented 13% of the antihypertensive drug costs whereas ARBs represented only 10%. ACEIs and CCBs are known to be among the most expensive antihypertensive drugs [41]. For instance, in Bahrain in one study, the cost of ACEIs represented two-thirds of the antihypertensive drug costs even though they were not the most widely used [38].

The US Association for Accessible Medicines reported that the CCB Norvasc (generic name: amlodipine) showed savings of USD 5.7 billion when the generic was substituted, drug savings that ranked among the top 10 [13]. To the best of our knowledge, no previous researchers in Kuwait aimed to impute the amount of savings that could be derived from substituting generic antihypertensive drugs for their branded originals in primary care centers. In the only study we are aware of, Cameron et al. estimated the amount of savings from substituting generics in a number of developing countries, and savings were lowest in Kuwait at 9% (int$ 44,785 = KD 13,778.11) [42]. The savings came from switching six drugs, two of which are antihypertensive drugs included in this study (captopril 25 mg and atenolol 50 mg), and the low savings were attributed to the low difference between the prices of the branded drugs and their equivalent generics [42]. In that study, generics cost more in Kuwait than in other countries, and innovator drugs were purchased from 1.2 to 32.9 times the international reference prices and generics from 0.1 to 22.2 times the international prices [42].

The low number of generics in Kuwait resulted in the absence of the competition that usually drives down drug prices. However, the MOH Pricing Department has been recently adjusting drug prices to account for drugs going off patent and taking into account prices in the region, and as such, it could be worthwhile to repeat this price study in the future to assess if higher savings could be achieved. Substituting a total of 890 antihypertensive drugs for 774 patients reduced total antihypertensive drug costs by 73.9%, with an average savings of KD 5.68. The 890 drugs substituted corresponded to 22 branded drugs. With an estimated 27.8% of the population living with elevated blood pressure, we estimate that Kuwait could achieve national cost savings of KD 81,328,010.4 per year for all patients with hypertension, and the sensitivity analysis showed that savings in the case of a lower prevalence of hypertension could still be significant.

Al Kanderi et al. 2009 reported in their study that there was a lack of essential antihypertensive drugs in primary health care clinics in Kuwait. According to the authors, MOH’s policy of confining expensive drugs to secondary and tertiary hospitals had likely driven overreliance on these facilities, where the increasing costs of antihypertensive drugs had made them unavailable in primary healthcare centers [43]. Higher costs and drug unavailability can lead to patient noncompliance with therapy and to uncontrolled hypertension, eventually increasing future health care expenditures due to complications [26]. Patients with hypertension often rely on primary health care clinics to regularly check their blood pressure and receive their monthly prescriptions, which is why it could be worthwhile to find a solution to this problem, such as switching to lower-priced generic drugs. Reducing the cost of antihypertensive drugs could increase their availability in primary health care clinics, reducing the pressure on secondary- and tertiary-level hospitals and increasing patient adherence to drug therapies.

Kuwait provides treatment free of charge to its citizens and at a subsidized cost for residents, so the government bears the cost of expensive drug brands. With the aging population and increases in non-communicable diseases, the price differences between branded and generic drugs should not be considered trivial. Even with low-cost medications, cost differences between generic and branded drugs in chronic diseases could translate into thousands of Kuwaiti dinars saved over a given patient’s life. Non-communicable diseases require recurrent and long-term medication services. In particular, cardiovascular diseases and hypertension take up a significant amount of total government health expenditures even as fewer persons might be affected as by other diseases. Cardiovascular diseases require frequent outpatient visits and expensive interventions, they result in intensive care unit admissions, and they are also associated with lengthier hospital stays and work absenteeism [23]. The MOH’s reliance on mostly innovator-branded drugs is unnecessary, and the potential savings from increasing the generic penetration in the Kuwaiti market are still untapped [30].

## Limitations

We encountered several challenges in the course of this study. Only three primary care centers were made available to be sampled, and all were from one governorate in Kuwait. Generalizing the current study findings will require a more representative sample from PHCCs across multiple regions of Kuwait. Moreover, we were forced to use the 2015 International Medical Products Price Guide when substituting generic drugs owing to the limited availability of generic drugs in Kuwait.

## Conclusion

Many countries have followed the recommendations of the WHO in promoting the use of generic drugs as a means of bringing down pharmaceutical expenditures and increasing access to medications. Kuwait has recently joined other countries in mandating prescription by generic name and increasing the availability of generics in the Kuwaiti pharmaceutical market, yet the generic prescription rate is still low. With the increasing prevalence of hypertension and its status as a lifelong illness came a necessity for decreasing its cost of treatment. With this study, we aimed to analyze the cost of drugs for managing hypertension in selected primary health care centers in Kuwait and to calculate the amount of savings that could be generated by substituting generic antihypertensive drugs for their branded equivalents. As we expected, only 71 generic drugs had been prescribed, making the generic prescription rate 4.0%, a rate the WHO considers very low. However, even at this low rate, substituting 890 antihypertensive drugs reduced total prescription costs by 56.23% and total antihypertensive drug costs by 73.91%, a promising amount that is in line with reports that prices of generics are anywhere between 10% and 90% less than those of originator drugs.

For this study, we estimated the national savings by assuming that all 1,306,600 patients with hypertension were treated in the same manner at the expense of the government, which carries the risk that we overestimated the savings. However, we did show that even for just one chronic condition, hypertension, the savings generated by substituting generic drugs for their originator brands could be significant enough to warrant policy changes. Barriers persist in Kuwait to the successful implementation of policies that encourage generic prescription, including the absence of national drug policy, the negative perception of the quality of generics, the heavy dependence on imported drugs, and the limited availability of generics in the Kuwaiti pharmaceutical market. These need to be addressed if Kuwait is to achieve a similar generic prescription rate to those reported by most developed countries.

## Data Availability

available upon request from

## Acknowledgments

Our deepest gratitude for the Primary Healthcare Division of MOH for acquiring the data that made this research possible. Our special thanks is offered to Dr. Donia AlBastaki, Head of the Drug Registration Department for providing information essential for the progress of this research. The authors would also like to thank Enago (www.enago.com) for the English language review.

## Author contributions

Concept and design: AO and SA were responsible for conceptualizing and designing the study. AO, SA, RA and MA^2^ were involved in acquisition of the data. AO & SA performed the analysis of the data. AO, SA and MA^1^ carried out the drafting of the manuscript. SA undertook critical revisions of the manuscript for important intellectual content.

## Declaring conflict of interest

None declared.

## Funding

No funding was received.

## Appendix

**Table 11:**
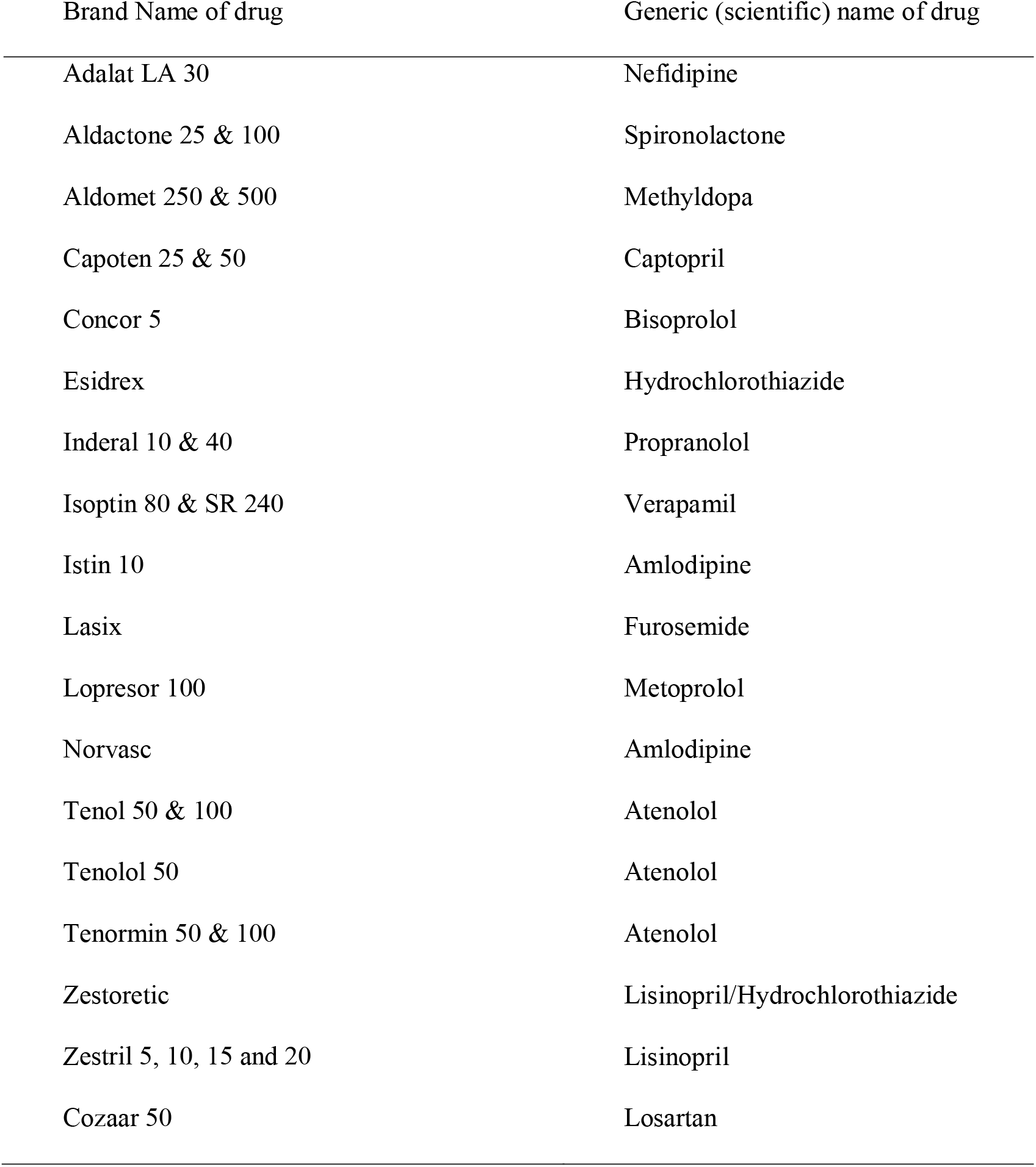
Substituted hypertension drugs

**Table 12:**
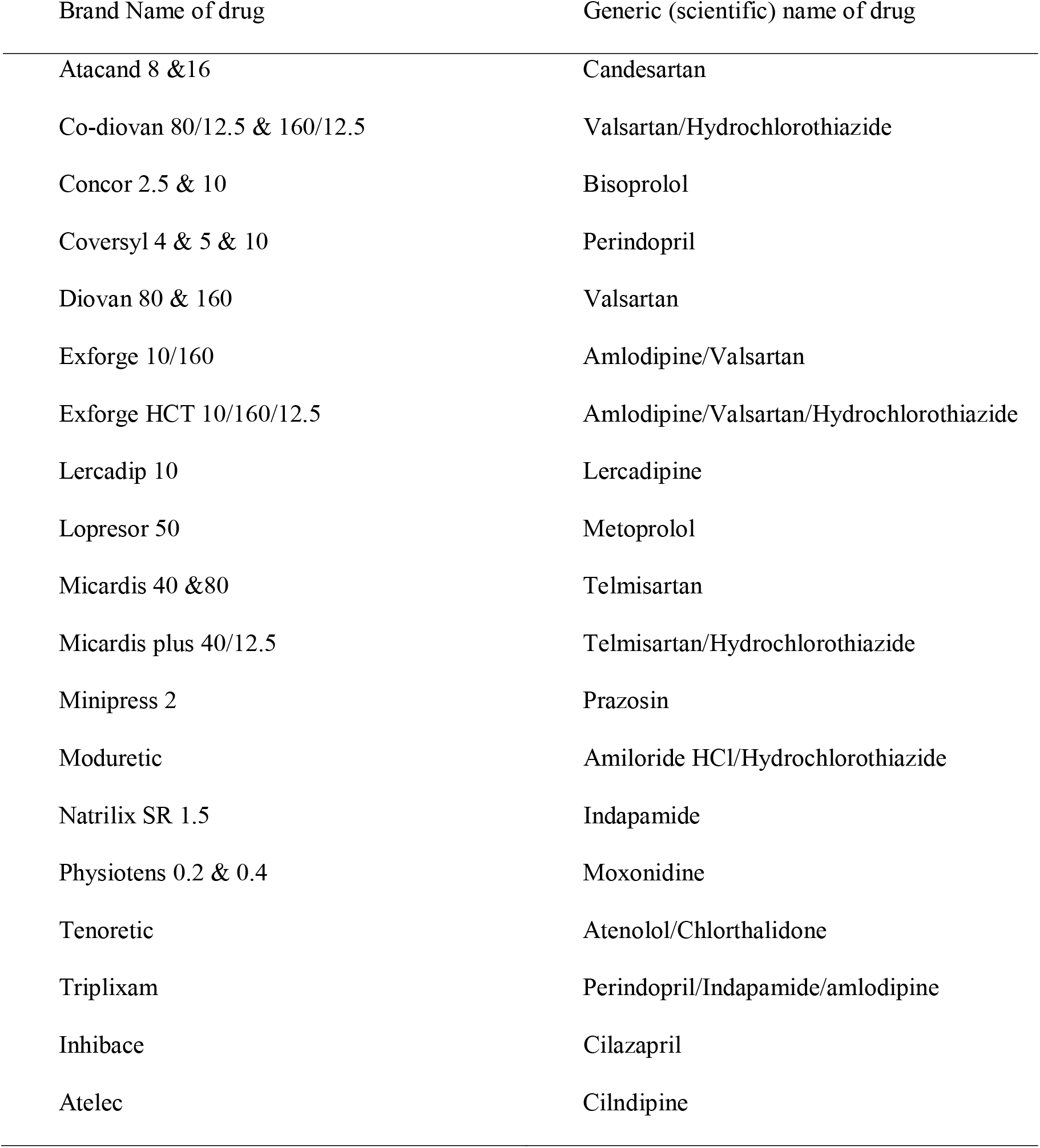
Unsubstituted hypertension drugs.

## Notes

### Competing Interest Statement

The authors have declared no competing interest.

### Funding Statement

no external funding was recieved

